# Beyond Anatomical Severity: Determinants of Health-Related Quality of Life and Transition Readiness in Adolescents with Congenital Heart Disease

**DOI:** 10.64898/2026.05.20.26353746

**Authors:** Mohammad Abed, Sandra Aiello, Navreet Gill, Rafael Alonso Gonzalez, Danielle Massarella, Rong Huang, Conall Thomas Morgan

## Abstract

**Background:** Improved survival of adolescents with congenital heart disease has shifted the focus to examine health-related quality of life and address challenges in transition to adult care. We aim to describe how congenital heart disease complexity, gender, number of interventions, and Fontan circulation may affect the health-related quality of life and transition readiness of adolescents with congenital heart disease.

**Methods:** We conducted a single-center cross-sectional study involving 536 patients aged 14 to 18 years old who attended a nurse-led, pediatric to adult care cardiac transition clinic, from 2020 to 2024. health-related quality of life was evaluated using the PedsQL^TM^ 4.0 Generic Core Scales and the PedsQL^TM^ 3.0 Cardiac Module. Patients were screened for anxiety and depression using the PHQ-9 and GAD-7. Transition readiness was assessed using the Transition-Q score.

**Results:** The median age of patients was 16 years old and 44% self-identified as female. PedsQL^TM^ 4.0 Generic had a median overall score of 77 (IQR 67–87), with no significant difference according to congenital heart disease severity. Female patients had significantly lower overall PedsQL^TM^ 4.0 score (p=0.028) and lower physical and emotional functioning scores (p=0.005, p<0.001, respectively) when compared to males. Physical functioning scores were lower amongst patients with Fontan circulation compared to non-Fontan patients (p=0.003), although overall PedsQL^TM^ 4.0 score and transition readiness scores were similar to those with complex biventricular congenital heart disease. Number of previous interventions were inversely associated with overall PedsQL^TM^ 4.0 score (p=0.036). Moderate to severe symptoms of depression or anxiety were reported in 30% of screened patients and were associated with significantly lower PedsQL^TM^ 4.0 scores (p<0.001). Transition readiness wa significantly lower in patients with moderate and complex compared to those with simple congenital heart disease (p<0.001). Transition readiness improved with repeat transition clinic visits (p=0.004) whereas PedsQL^TM^ 4.0 score did not change significantly.

**Conclusion:** In this large cohort of adolescents with congenital heart disease, health-related quality of life was lower than population norms. Female gender, higher interventional burden, and anxiety or depressive symptoms are associated with lower health-related quality of life scores rather than anatomical severity or Fontan physiology. Transition readiness was lower in complex disease; it has improved with a structured, nurse-led transition clinic, demonstrating modifiability. Consequently, adolescent congenital heart disease care requires a multidisciplinary approach including psychosocial screening, especially for high-risk groups, and structured transition planning to improve long-term outcomes.

## INTRODUCTION

Progress in surgical, interventional, and medical therapies for children with congenital heart disease (CHD) has led to considerable improvements in survival across all disease severities, including patients with a Fontan circulation.^1^ With decreasing mortality, there is an increased focus on the long-term morbidities that can affect survivors, including physical, psychosocial, and neurodevelopmental impairments. These morbidities can impact their health-related quality of life (HRQOL).^2^ HRQOL is broadly defined as the impact of a specific illness, medical therapy, or health services policy on a patient’s ability to function in various life contexts and to derive personal satisfaction from physical, psychological, and social functioning.^3,2^ Assessing HRQOL is important in adolescents with CHD, as it may be covertly impacted by the type of heart disease, their interventional history and their residual lesions. This can, however, be challenging, as parent proxy reports are often relied on and may differ significantly from the patient’s perspective.^3^ Increased life expectancy among adolescents with congenital heart disease (CHD) has shifted focus to comorbidities such as heart failure, arrhythmia, and the need for repeat interventions, highlighting the necessity of effective transition to adult care. HRQOL, mental health and transition readiness are closely interrelated, as diminished physical or psychosocial well-being may hinder an adolescent’s capacity to manage their disease independently.^4^ Many adolescents possess limited knowledge of their medical condition, primarily because parents typically oversee all aspects of disease management, including information acquisition, medication administration, follow-up arrangements, and decision-making. ^5^

The aim of our study was to describe how HRQOL among adolescents with CHD is affected by CHD severity, gender, interventional history, presence of Fontan circulation, and mental health burdens such as anxiety and depression. We hypothesized that greater CHD severity and the presence of Fontan circulation would negatively impact HRQOL. As a secondary aim, we assessed the effect of a nurse-led transition clinic on patients’ transition readiness and HRQOL.

## METHODS

### Study Design and Patient Population

A single-center, cross-sectional study was performed at a tertiary-level pediatric hospital in patients with CHD. Patients aged 14-18 years old who attended nurse-led cardiac transition clinic between January 2020 and December 2024 were included in the study. Patients were invited to attend the transition clinic alongside their clinical visits. Patients were excluded if they were unable to complete the questionnaires independently in English.

### Study measures

To evaluate HRQOL, patients completed two questionnaires administered via REDCap prior to attending the cardiac transition clinic. The first was the Pediatric Quality of Life Inventory^TM^ 4.0 Generic Core Scales (PedsQL^TM^ 4.0), a validated 23-item generic measure of HRQOL.^6^ It is comprised of four subscales: physical functioning, emotional functioning, social functioning, and school functioning. Scores from a 5-point Likert scale, ranging from 0 (Never) to 4 (Almost always), were transformed to generate a scale from 0 to 100, where higher scores indicate better quality of life. The second questionnaire was the Pediatric Quality of Life Inventory^TM^ 3.0 Cardiac Module,^6^ a 27-item assessment module for adolescents with heart disease. This module includes six domains: Heart Problems and Treatment, Treatment II, perceived physical appearance, treatment anxiety, cognitive problems, and communication. Responses are on a 5-point Likert scale, with higher scores per domain indicating better HRQOL and fewer problems or symptoms.

If the medical team identified clinical suspicion or concerns regarding anxiety or depression in a patient, the Patient Health Questionnaire-9 (PHQ-9) and General Anxiety Disorder-7 (GAD-7) questionnaires were administered by the nursing team. The PHQ-9, a 10-item specific instrument designed to screen for major depressive disorder, classifies depression severity as minimal (0–4), mild (5–9), moderate (10–14), moderately-severe (15–19), and severe (20–27).^7^ The GAD-7, is a 7-item specific instrument designed to screen for generalized anxiety disorder, categorizing anxiety severity as minimal (0–4), mild (5–9), moderate (10–14), and severe (15–21).^8^ Both questionnaires are widely used tools, with higher scores showing more severe symptoms.

To evaluate the impact of a nurse-led transition clinic on transition readiness and health-related quality of life, patients who attended the clinic more than once were assessed longitudinally. We used the Transition-Q score.^9^ Transition-Q contains 14 items that assess transition readiness, including knowledge, self-management, communication, and responsibility for one’s health. Each item is scored on a three-point response scale reflecting the individual’s current level of preparedness or independence. Higher cumulative scores indicate greater readiness for transition to adult-oriented care.

Patient clinical information was collected from the electronic patient record, including self-reported gender, CHD diagnosis, and number of prior cardiac catheterizations and surgical interventions. CHD severity was classified as simple, moderate, or complex according to the American College of Cardiology/American Heart Association adult congenital heart disease guidelines.^10^

## ETHICS

The study was approved by the Research Ethics Board of The Hospital for Sick Children (study number 1000081179).

## STATISTICAL ANALYSIS

Continuous variables were reported as medians with interquartile ranges (IQR), while categorical variables (dichotomous or polytomous) were summarized as frequencies and percentages. Between-group differences in continuous variables were evaluated using the Wilcoxon rank-sum test (two groups) or Kruskal-Wallis test (three or more groups), and differences in categorical variables were assessed using Fisher’s exact test.

In the presence of statistical significance in between-group differences for continuous variables, a pairwise Wilcoxon test was conducted to determine which specific group differs from each other. Adjusted P values were reported using Bonferroni methods to control the family-wise error rate in multiple comparisons.

## RESULTS

### Patient Characteristics

Five hundred thirty-six adolescents were included in the study (Table 1). Females accounted for 44% of the cohort (n=234), and the median age was 16 years (IQR: 15-17 years). Simple CHD lesions accounted for 6% (n=33), moderate for 58% (n=310) and complex 36% (n=193) of the entire cohort. Age and gender distributions were similar across CHD severity levels. Those with greater disease complexity were more often required two or more interventions (simple: 9%; moderate: 44%; severe: 84%; p<0.001). There were 65 patients with Fontan circulation. The overall median PedsQL^TM^4.0 score for the entire cohort was 77 (IQR 67-87). For the PedsQL^TM^ 3.0 Cardiac Module, one patient’s score was not computed because more than 50% of the items were missing. Among the participants, 109 adolescents who were taking medication at the time of the study completed the drug-related treatment subscale. The remaining five subscales were relevant to all participants (n=535).

**Table 1.** Patient Characteristics. Table 1 summarizes baseline patient characteristics stratified by Sex, CHD complexity, number of previous intervention and presence of Fontan circulation.

| Variable | Overall<br>(n=536) | Simple<br>(n=33) | Moderate<br>(n=310) | Complex<br>(n=193) | P-value |
| --- | --- | --- | --- | --- | --- |
| Female, n (%) | 234 (43.7%) | 17 (51.5%) | 142 (45.8%) | 75 (38.9%) | 0.194 |
| Age (years), median (IQR) | 16 (15–17) | 17 (16–17) | 16 (15–17) | 16 (15–17) | 0.103 |
| Number of Interventions, n (%) |  |  |  |  | <0.001 |
| 0 | 37 (6.9%) | 11 (33.3%) | 23 (7.4%) | 3 (1.6%) |  |
| 1 | 198 (36.9%) | 19 (57.6%) | 151 (48.7%) | 28 (14.5%) |  |
| 2 | 99 (18.5%) | 2 (6.1%) | 59 (19.0%) | 38 (19.7%) |  |
| 3 | 57 (10.6%) | 0 (0.0%) | 37 (11.9%) | 20 (10.4%) |  |
| 4 | 41 (7.6%) | 1 (3.0%) | 22 (7.1%) | 18 (9.3%) |  |
| 5 | 34 (6.3%) | 0 (0.0%) | 5 (1.6%) | 29 (15.0%) |  |
| 6 | 25 (4.7%) | 0 (0.0%) | 7 (2.3%) | 18 (9.3%) |  |
| 7 | 14 (2.6%) | 0 (0.0%) | 2 (0.6%) | 12 (6.2%) |  |
| 8–14 | 31 (5.8%) | 0 (0.0%) | 4 (1.3%) | 27 (14.0%) |  |

### Effect of Gender

Female patients scored significantly lower than males on the overall PedsQL^TM^4.0 score [75 (64-87) vs 79 (69-88), p=0.028] (Table 2). Females have scored lower than males in the subdomains of physical functioning [84 (72-94) vs 88 (78-97), p=0.005] and emotional functioning [70 (55-85) vs 75 (65-90), p<0.001]. Within the PedsQL^TM^ 3.0 Cardiac Module subdomains, females scored significantly less than males in perceived physical appearance [75 (50-92) vs 83 (67-100), p<0.001] and treatment anxiety [81 (63-100) vs 94 (69-100), p=0.015] with a trend towards lower scores in the communication subdomain [67 (50-83) vs 75 (58-92), p=0.07]. No significant differences were observed in the subdomains of cognitive problems and communication.

**Table 2.** PedsQL^TM^4 and PedsQL Cardiac Module Subdomains Scores according to Gender. Table 2 summarizes the PedsQL^TM^4.0 scores and PedsQL Cardiac Module Subdomains Scores by gender among adolescents with congenital heart disease, reported as median and interquartile range.

| **Variables** | **Female N** | **Female Median [IQR]** | **Male N** | **Male Median [IQR]** | **P-value** |
| --- | --- | --- | --- | --- | --- |
| Physical function | 234 | 84 (72 – 94) | 302 | 88 (76 – 97) | 0.005 |
| Emotional function | 234 | 70 (55 – 85) | 302 | 75 (65 – 90) | <0.001 |
| Social function | 234 | 85 (70 – 100) | 302 | 90 (75 – 100) | 0.183 |
| School function | 234 | 65 (55 – 80) | 302 | 65 (50 – 80) | 0.815 |
| **PedsQL™4 total score** | **234** | **75 (64 – 87)** | **302** | **79 (69 – 88)** | **0.028** |
| Heart problems and treatment | 234 | 79 (64 – 86) | 302 | 79 (68 – 89) | 0.015 |
| Treatment II | 37 | 90 (85 – 95) | 72 | 95 (90 – 95) | 0.47 |
| Perceived physical appearance | 234 | 75 (50 – 92) | 301 | 87 (66 – 100) | <0.001 |
| Treatment anxiety | 234 | 81 (63 – 100) | 301 | 94 (69 – 100) | 0.015 |
| Cognitive problems | 234 | 65 (45 – 90) | 301 | 71 (50 – 90) | 0.012 |
| Communication | 234 | 67 (50 – 83) | 301 | 75 (58 – 92) | 0.075 |

### Fontan circulation

Compared with patients without Fontan circulation, a trend toward a lower overall PedsQL^TM^4.0 score was observed [73 (64-84) vs 78 (68-88), p=0.06] (Table 3). Fontan patients had a lower physical functioning subdomain score than non-Fontan patients [81 (72-91) vs 88 (75-97), p=0.003]. No other subdomain in either tool was significantly different. When comparing Fontan patients to patients with other complex CHD, no differences were observed in overall PedsQL^TM^4.0 scores. The only subdomain that was significantly lower in patients with Fontan compared to patients with other complex CHD was physical functioning [81 (72-91) vs 88 (75-94), p=0.037] (Table S1). In terms of transition readiness, there was no difference between Fontan patients and non-Fontan patients [14 (11-17) vs 14 (11-18), p=0.57] or Fontan and other forms of complex CHD [14 (11-17) vs 15 (12-17), p=0.57]. (Table S2)

**Table 3.**
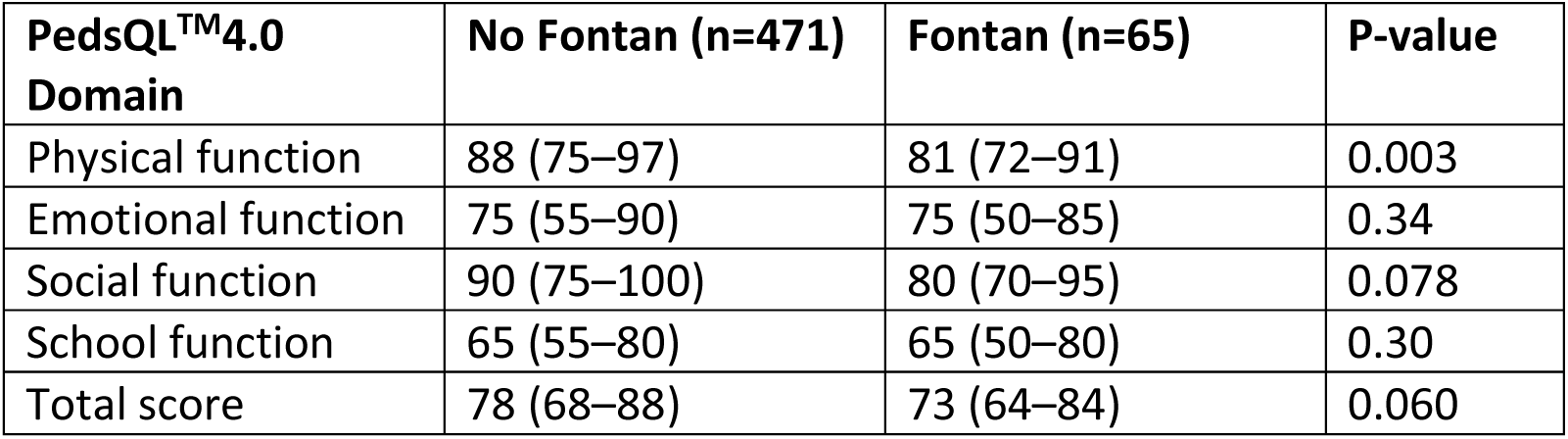
PedsQL^TM^4.0 scores by Fontan Status. Table 3 compares PedsQL^TM^4 scores between adolescents with and without Fontan circulation.

| **PedsQL™4.0 Domain** | **No Fontan (n=471)** | **Fontan (n=65)** | **P-value** |
| --- | --- | --- | --- |
| Physical function | 88 (75–97) | 81 (72–91) | 0.003 |
| Emotional function | 75 (55–90) | 75 (50–85) | 0.34 |
| Social function | 90 (75–100) | 80 (70–95) | 0.078 |
| School function | 65 (55–80) | 65 (50–80) | 0.30 |
| Total score | 78 (68–88) | 73 (64–84) | 0.060 |

### Number of Interventions

The number of interventions is shown in Table 1. Overall PedsQL^TM^4.0 score was inversely proportional to the number of interventions (r=-0.090, p=0.036) (Table S3). The subdomains of physical functioning (r=-0.132, p=0.002), social functioning (r=-0.102, p=0.018) and heart problems and treatment (r=-0.143, p<0.001) negatively correlated with number of interventions.

### CHD severity

CHD severity was not significantly associated with a lower PedsQL^TM^4.0 score (Table 4). In the subdomain of physical functioning, scores were significantly lower with increasing CHD severity [simple 91 (78-100), moderate 88 (75-97), complex 84 (72-94), p = 0.031]. There was no significant difference in the subdomains of emotional, social or school functioning related to CHD severity. In the cardiac module subdomains of heart problems and treatment, scores were significantly lower with increasing CHD severity [simple 82 (75-93), moderate 79 (68-89), complex 79 (64-89), p=0.05]. No statistically significant association was observed among the other subdomains. Transition-Q scores were significantly lower in patients with moderate or complex CHD than in those with simple CHD [15 (11-17) vs 17 (15-20), p<0.001].

**Table 4.** PedsQL Cardiac Module Scores according to CHD Complexity. Table 4 presents the PedsQL^TM^ 3.0 Cardiac Module subdomains scores according to CHD complexity groups.

| **PedsQL Cardiac**<br>**Module Domain** | **Overall**<br>**(n=536)** | **Simple (n=33)** | **Moderate**<br>**(n=310)** | **Complex**<br>**(n=193)** | **P-value** |
| --- | --- | --- | --- | --- | --- |
| Heart problems and treatment | 79 (68–89) | 82 (75–93) | 79 (68–89) | 79 (64–89) | 0.050 |
| Treatment II | 90 (85–95) | 100 (95–100) | 95 (90–95) | 90 (85–95) | 0.34 |
| Perceived physical appearance | 83 (58–100) | 75 (58–92) | 83 (58–92) | 83 (67–100) | 0.30 |
| Treatment anxiety | 88 (69–100) | 94 (75–100) | 88 (69–100) | 88 (69–100) | 0.64 |
| Cognitive problems | 65 (50–80) | 65 (50–85) | 65 (50–80) | 65 (50–80) | 0.80 |
| Communication | 75 (58–92) | 75 (58–92) | 67 (50–92) | 75 (58–92) | 0.68 |

### Presence of anxiety and/or depressive symptoms

A total of 106 (20%) patients were administered the PHQ-9 and GAD-7 questionnaires. Overall, moderate or greater depressive symptoms were seen in 31 patients (29%) and moderate or greater anxiety symptoms in 33 patients (31%). There was no observed difference in the frequency of moderate or greater depression or anxiety symptoms with increasing CHD severity. Comparison of patients with Fontan circulation to those without, there was no significant difference in frequency of moderate or greater depressive symptoms [23% (n=3) vs 30% (n=28), p=0.93] or moderate or greater anxiety symptoms [31% (n=4) vs 31% (n=29), p=1.00]. (Table S4).

Moderate or severe symptoms of depression were associated with a statistically significant lower overall PedsQL^TM^4.0 score [59 (49-77) vs 74 (70-84), p<0.001]. All scores were similarly lower across all subdomains in both the PedsQL^TM^4.0 and the cardiac module, except for Treatment II and treatment anxiety. Moderate or greater symptoms of anxiety, compared with none or mild symptoms, were similarly associated with a significantly lower PedsQL^TM^4.0 overall score [66 (50-76) vs 74 (68-85), p=0.001]. Participants with moderate or greater anxiety had lower scores in all the subdomains except for heart problems, treatment and Treatment II. Transition readiness scores were similar between patients with moderate or greater depression compared with no or mild depression [14 (12-18) vs 15 (11-18), p=0.5] (Table S5) and similarly in patients with moderate to severe anxiety symptoms compared with no or mild symptoms [14 (11-16) vs 15 (12-19), p=0.129].

### Transition readiness and impact of the nursing-led transition clinic

Patients with simple CHD demonstrated higher transition readiness scores than those with moderate or complex CHD (median 17 versus 14 and 15, respectively; P < 0.001).(Table 5) In the Fontan patient group, transition readiness scores were lower than those observed in the complex CHD subgroup; however, this difference was not statistically significant (median 14 versus 15, P = 0.57).(Table S2) Among 118 patients who attended the transition clinic twice, the transition readiness score increased significantly (14 vs 16, P = 0.004) (Figure 1). PedsQL^TM^4.0 score and PedsQL^TM^ cardiac module subdomains scores did not change significantly following the second transition clinic visit. (Table S2).

**Figure 1.**
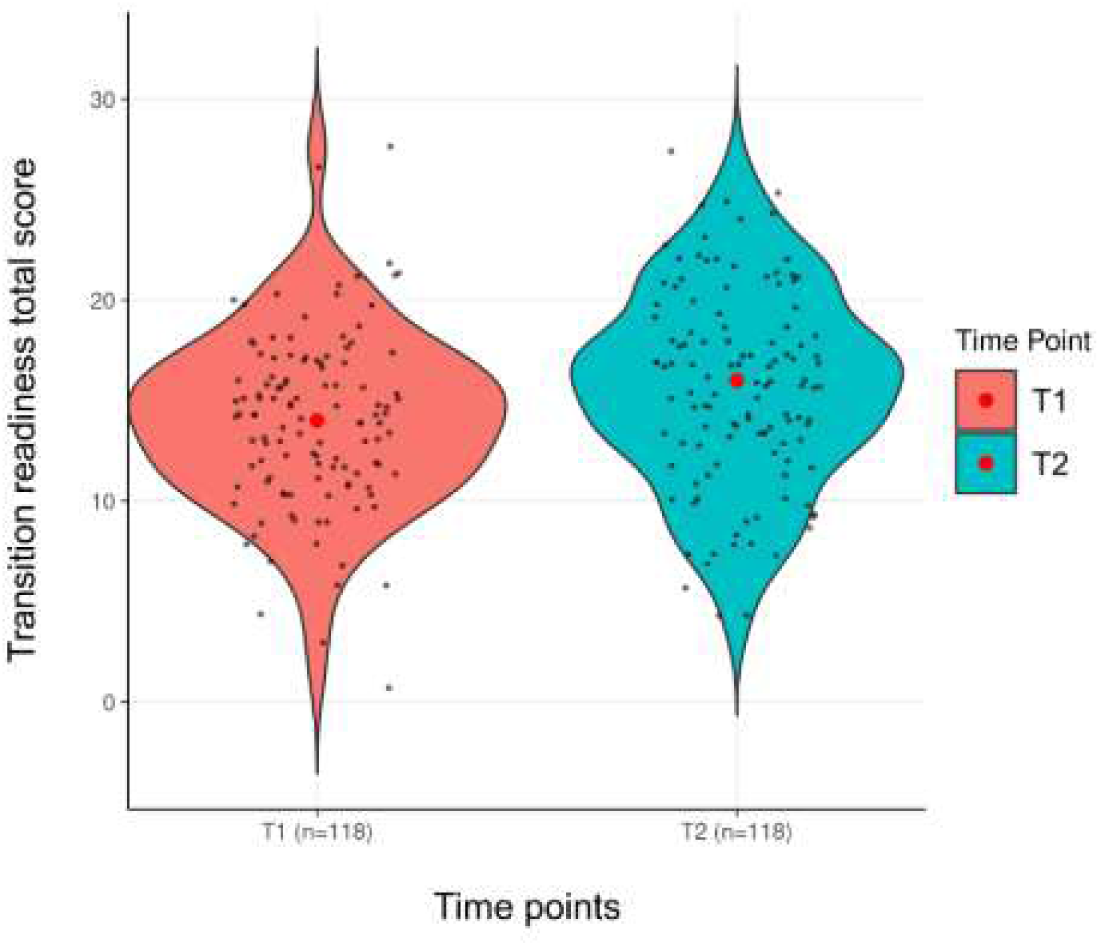
Transition readiness score changes over time. Transition readiness total scores were assessed at the first and second transition clinic visits (n = 118). Median scores increased from 14 (IQR 11–17) at the first visit to 16 (IQR 13–19) at the second visit (P = 0.004).

**Table 5.** Transition Q score by CHD severity. Table 5 Transition-readiness scores among adolescents with CHD by complexity. Data are presented as n (%) or median (IQR).

| **Variable** | **Overall (n = 536)** | **Simple (n = 33)** | **Moderate (n = 310)** | **Complex (n = 193)** | **P-value** |
| --- | --- | --- | --- | --- | --- |
| **I answer a doctor's or nurse's questions** |  |  |  |  | 0.42 |
| Never | 9 (1.7%) | 0 (0.0%) | 5 (1.6%) | 4 (2.1%) |  |
| Sometimes | 192 (35.8%) | 8 (24.2%) | 108 (34.8%) | 76 (39.4%) |  |
| Always | 335 (62.5%) | 25 (75.8%) | 197 (63.5%) | 113 (58.5%) |  |
| **I help to make decisions about my health** |  |  |  |  | 0.34 |
| Never | 32 (6.0%) | 1 (3.0%) | 20 (6.5%) | 11 (5.7%) |  |
| Sometimes | 242 (45.1%) | 10 (30.3%) | 140 (45.2%) | 92 (47.7%) |  |
| Always | 262 (48.9%) | 22 (66.7%) | 150 (48.4%) | 90 (46.6%) |  |
| **I am in charge of taking any medicine that I need** |  |  |  |  | 0.34 |
| Never | 63 (11.8%) | 1 (3.0%) | 37 (11.9%) | 25 (13.0%) |  |
| Sometimes | 165 (30.8%) | 9 (27.3%) | 102 (32.9%) | 54 (28.0%) |  |
| Always | 308 (57.5%) | 23 (69.7%) | 171 (55.2%) | 114 (59.1%) |  |
| **I talk to a doctor or nurse when I have health concerns** |  |  |  |  | 0.76 |
| Never | 51 (9.5%) | 2 (6.1%) | 30 (9.7%) | 19 (9.8%) |  |
| Sometimes | 239 (44.6%) | 12 (36.4%) | 142 (45.8%) | 85 (44.0%) |  |
| Always | 246 (45.9%) | 19 (57.6%) | 138 (44.5%) | 89 (46.1%) |  |
| **I look for an answer when I have a question about my health** |  |  |  |  | 0.85 |
| Never | 41 (7.6%) | 1 (3.0%) | 26 (8.4%) | 14 (7.3%) |  |
| Sometimes | 195 (36.4%) | 11 (33.3%) | 115 (37.1%) | 69 (35.8%) |  |
| Always | 300 (56.0%) | 21 (63.6%) | 169 (54.5%) | 110 (57.0%) |  |
| **I talk about my health condition to people when I need to** |  |  |  |  | 0.022 |
| Never | 45 (8.4%) | 2 (6.1%) | 30 (9.7%) | 13 (6.7%) |  |
| Sometimes | 183 (34.1%) | 4 (12.1%) | 113 (36.5%) | 66 (34.2%) |  |
| Always | 308 (57.5%) | 27 (81.8%) | 167 (53.9%) | 114 (59.1%) |  |
| **I ask the doctor or nurse questions** |  |  |  |  | 0.012 |
| Never | 60 (11.2%) | 1 (3.0%) | 40 (12.9%) | 19 (9.8%) |  |
| Sometimes | 297 (55.4%) | 12 (36.4%) | 177 (57.1%) | 108 (56.0%) |  |
| Always | 179 (33.4%) | 20 (60.6%) | 93 (30.0%) | 66 (34.2%) |  |
| **I speak to the doctor instead of my parent(s) speaking for me** |  |  |  |  | 0.56 |
| Never | 85 (15.9%) | 3 (9.1%) | 52 (16.8%) | 30 (15.5%) |  |
| Sometimes | 358 (66.8%) | 22 (66.7%) | 202 (65.2%) | 134 (69.4%) |  |
| Always | 93 (17.4%) | 8 (24.2%) | 56 (18.1%) | 29 (15.0%) |  |
| **I summarize my medical history when I am asked to** |  |  |  |  | 0.71 |
| Never | 163 (30.4%) | 7 (21.2%) | 92 (29.7%) | 64 (33.2%) |  |
| Sometimes | 217 (40.5%) | 15 (45.5%) | 128 (41.3%) | 74 (38.3%) |  |
| Always | 156 (29.1%) | 11 (33.3%) | 90 (29.0%) | 55 (28.5%) |  |
| **I contact a doctor when I need to** |  |  |  |  | 0.004 |
| Never | 193 (36.0%) | 3 (9.1%) | 115 (37.1%) | 75 (38.9%) |  |
| Sometimes | 165 (30.8%) | 16 (48.5%) | 99 (31.9%) | 50 (25.9%) |  |
| Always | 178 (33.2%) | 14 (42.4%) | 96 (31.0%) | 68 (35.2%) |  |
| **I see the doctor or nurse on my own during an appointment** |  |  |  |  | 0.71 |
| Never | 300 (56.0%) | 15 (45.5%) | 175 (56.5%) | 110 (57.0%) |  |
| Sometimes | 193 (36.0%) | 14 (42.4%) | 110 (35.5%) | 69 (35.8%) |  |
| Always | 43 (8.0%) | 4 (12.1%) | 25 (8.1%) | 14 (7.3%) |  |
| **I drop off or pick up my prescriptions when I need medicine** |  |  |  |  | <0.001 |
| Never | 353 (65.9%) | 10 (30.3%) | 212 (68.4%) | 131 (67.9%) |  |
| Sometimes | 112 (20.9%) | 11 (33.3%) | 59 (19.0%) | 42 (21.8%) |  |
| Always | 71 (13.2%) | 12 (36.4%) | 39 (12.6%) | 20 (10.4%) |  |
| **I travel on my own to a doctor's appointment** |  |  |  |  | 0.182 |
| Never | 463 (86.4%) | 25 (75.8%) | 268 (86.5%) | 170 (88.1%) |  |
| Sometimes | 58 (10.8%) | 7 (21.2%) | 35 (11.3%) | 16 (8.3%) |  |
| Always | 15 (2.8%) | 1 (3.0%) | 7 (2.3%) | 7 (3.6%) |  |
| **I book my own doctor's appointments** |  |  |  |  | 0.27 |
| Never | 456 (85.1%) | 24 (72.7%) | 267 (86.1%) | 165 (85.5%) |  |
| Sometimes | 68 (12.7%) | 8 (24.2%) | 37 (11.9%) | 23 (11.9%) |  |
| Always | 12 (2.2%) | 1 (3.0%) | 6 (1.9%) | 5 (2.6%) |  |
| **Transition readiness total score** | 14 (11–17) | 17 (15–20) | 14 (11–17) | 15 (11–17) | <0.001 |

## DISCUSSION

### HRQOL in adolescents with CHD compared with healthy peers and adolescents with other chronic diseases

The typically reported overall PedsQL^TM^4.0 score for healthy adolescents is 84, with lower scores in females and the lowest scores in low-income countries.^11^ In our study, the overall PedsQL^TM^4.0 score was 77, lower than that of healthy adolescents.^12^ This is consistent with previously published reports for patients with CHD.^13^ In adults with CHD, as well as other pediatric chronic illnesses such as cancer, studies have shown that patients usually report lower quality of life scores than the general population.^14,13^ Newly diagnosed adolescents with cancer on active treatment report lower HRQOL, while those in long-term remission after more than 1 year fare better.^15^ Though CHD lacks clearly defined treatment phases like those in oncology, the two illnesses share similarities in terms of chronicity and cumulative intervention burden. Similarly, across other chronic pediatric conditions, children with rheumatologic disorders and those with type 1 and type 2 diabetes demonstrate reduced HRQOL compared with healthy peers, with reported mean scores of 74 and 80, respectively.^16^ Collectively, these comparisons suggest that adolescents with CHD experience a degree of HRQOL impairment comparable to other chronic pediatric conditions, reinforcing the pervasive impact of chronic disease.

Our cohort showed that adolescents with more cardiac interventions reported significantly lower PedsQL^TM^4.0 scores, particularly in the physical and social functioning domains, indicating that a high interventional burden may intensify perceived differences between patients and healthy peers. O’Connor et al.have previously described this: the higher the burden of procedures, the greater the negative impact on overall well-being.^2^ Paramedical support, such as social work and/or psychology, may be beneficial for these adolescents.

There is some heterogeneity in the reported literature on HRQOL in adolescents with CHD. Some studies report lower HRQOL scores compared to healthy peers, while others report comparable or even better perceived QoL among CHD patients.^17^ This variation highlights the influence of socioeconomic, psychosocial and contextual factors, such as coping strategies and social support, in addition to simple lesion anatomy.^13^ While we did not see lower PedsQL^TM^4.0 scores with increasing disease severity, other studies have. Varni et al. found a clear relationship between more complex cardiac disease and lower HRQOL. It is possible that children with chronic illnesses, including those with cardiac conditions, develop a unique mechanism of adaptation secondary to their experiences. Growing up with CHD requires managing medical treatment and may involve activity restrictions. Since these children have not known a different way of living, they may see and value life in their own way. Their sense of quality of life is often influenced by their abilities and limitations.^14,13^

### Fontan circulation

Adult patients with Fontan circulation tend to report lower HRQOL scores compared to healthy populations and in patients with repaired biventricular lesions, especially in the domains of physical and social functioning.^18^ Suggesting that the long-term physiological and psychosocial impact of single ventricle circulation begins early and persists across the lifespan.^19^

In our study, patients with Fontan circulation demonstrated HRQOL scores comparable to those of adolescents with biventricular circulations. However, Fontan patients showed significantly lower scores in the domains of physical functioning, heart problems and treatment. Several factors may account for the similarity in overall HRQOL between Fontan and non-Fontan adolescents with CHD. First, the Fontan population is heterogeneous, and adolescents who reach this stage of palliation and attend outpatient follow-up are often those with relatively stable physiology and fewer complications. In this study, we did not stratify Fontan patients based on clinical status, such as ventricular function, arrhythmia burden, exercise capacity, or Fontan-related complications. As a result, our cohort may have included mostly clinically stable Fontan patients whose daily functioning and perceived quality of life are comparable to those of peers with other repaired congenital heart conditions. Second, Fontan adolescent is a period during which many Fontan-related complications have not yet fully developed. Unlike adult cohorts, where Fontan failure, arrhythmia, liver disease, and reduced exercise capacity become more common. Adolescents often retain a relatively preserved functional status. Therefore, the psychosocial and physical effects of the Fontan circulation observed in adult studies may not yet be fully evident during adolescence.

Overall, these findings suggest that the impact of Fontan physiology on perceived quality of life may change over time, with adolescents maintaining HRQOL similar to peers with other congenital heart lesions, while the increased physical and psychosocial challenges seen in adults may appear later in the disease course.

### Effect of gender

The observed gender-based differences in PedsQL^TM^ 4.0 and PedsQL^TM^ 3.0 cardiac module scores highlight a consistent trend of lower HRQOL scores among female adolescents with CHD, in contrast to some prior literature where no differences were witnessed when comparing both gender subgroups.^20^ Females reported significantly lower scores across multiple domains, including physical and emotional functioning, cognitive problems, treatment anxiety, and perceived physical appearance. These findings are consistent with a prior study by Cousino et al., suggesting that female adolescents with chronic illnesses may experience greater physical burden, which is newly described as related to body image concerns.^21^ The lower cardiac module scores among females may also reflect heightened sensitivity to symptoms, treatment-related stress, or social pressures. Further investigation of these disparities is necessary to identify modifiable factors and guide the development of gender-specific support strategies in long-term cardiac care. These findings can help caregivers become more aware and facilitate the identification of patients who would benefit from targeted interventions or referral to relevant professional services.

### Anxiety and depression

In young adults with complex CHD, the prevalence of anxiety and depression is estimated to range between 30 to 40%.^22^ Miles et al. described that over one third of children with congenital heart disease experience some form of diagnosable mental health condition based on a nationwide Danish population cohort.^23^ Recently, the American Heart Association emphasized that adolescents with CHD face unique developmental and disease-related stressors, including academic challenges, physical limitations, and social anxiety, all of which contribute to poorer quality of life, particularly when depressive symptoms and loneliness are present.^24^ In our study, 29% reported at least moderate depression, and 31% reported at least moderate anxiety symptoms. These figures exceed the reported anxiety prevalence among healthy adolescents aged 11 to 17 years old in the United States (6% to 11%), as described by Ghandour et al.^25^ Though these figures did align with rates seen in children with other chronic conditions such as end-stage renal disease, asthma, obesity, and diabetes mellitus.^3,26^

Our study shows that adolescents with moderate to severe anxiety or depression symptoms report lower HRQOL scores compared to those with minimal or mild symptoms. Similarly, Cohen et al. showed that a depressed mood in adolescents with heart disease led to lower HRQOL.^27^ This clearly highlights that the degree of anxiety and depression is inversely associated with quality of life. This is clinically relevant, as validated screening tools for anxiety and depression are widely available and can be feasibly integrated into routine cardiology practice. Subgroup analysis of anxiety and depression prevalence based on CHD severity showed no significant differences in reported anxiety or depressive symptoms relative to CHD complexity. This might indicate that even adolescents with simple CHD lesions may experience psychological distress, probably due to the cumulative impact of repeated hospital visits and ongoing medical investigations.

Recent data from the Fontan Network Outcomes by Glenn et al. showed that anxiety and mood disorders increase significantly with age, affecting over 30% of the registry.^28^ In our study, 31% of Fontan patients were clinically suspected of having anxiety or depression, which aligns with the overall prevalence of mental health burden in this population.

### Transition readiness and impact of a transition clinic

Transition readiness in our cohort demonstrates several clinically relevant patterns that are consistent with prior literature. Patients with simple CHD had higher readiness scores than those with moderate or highly complex disease. This finding suggests that complex disease make it more challenging to acquire sufficient understanding of the disease and self-management skills. These results align with previous work by Stewart et al., who emphasized that transition programs should focus on improving patient knowledge and self-management competencies, particularly in more complex populations.^29^ Our findings underscore the positive impact that structured transition clinic exposure has. Adolescents who attended serial transition clinic visits showed significant improvements in transition readiness score. These results indicate that clinic-based, nurse-led transition interventions can quickly enhance readiness scores, as Mackie et al. have previously demonstrated.^30^

## CONCLUSION

In this large single-center cohort of adolescents with congenital heart disease, HRQOL was lower than population norms and was more affected by patient-specific and psychosocial factors rather than by anatomical disease severity. Female sex, higher interventional burden, and anxiety or depressive symptoms were consistently linked to lower HRQOL. In contrast, CHD severity and Fontan physiology had less impact. Transition readiness was lower in patients with more complex disease but improved with repeated participation in a structured, nurse-led transition clinic, demonstrating that this outcome is modifiable. These findings highlight the need for a comprehensive, multidisciplinary approach to adolescent CHD care that goes beyond anatomical classification. Care should involve psychosocial screening, focused support for high-risk groups, and planned transition steps to help improve long-term outcomes and encourage adherence.

## STRENGTHS AND LIMITATIONS

This research uses well-defined cohort and validated instruments, including cardiac-specific modules, to assess quality of life, anxiety, and depressive symptoms. The questionnaires and outcome data were based on patient-reported measures instead of parent-proxy reports, this approach allows for a direct assessment of the patients’ own experiences and perceptions. The survey was distributed 1 week in advance of the clinic visit to give patients sufficient time to complete it thoroughly. Although the cohort was conducted from a single tertiary care center in North America, which may limit the generalizability of the findings, the study population was nonetheless ethnically and culturally diverse, reflecting the center’s catchment area’s multicultural composition. Transition Q scores were assessed at two separate clinic visits, which strengthened the reliability and validity of the intervention.

This study has several limitations. A retrospective analysis without a longitudinal design cannot establish causal relationships. The median age of the cohort was 16 years, which restricts the generalizability of these findings to older patients with CHD who may experience lower HRQOL as morbidities increase with age, particularly within the Fontan subgroup. Furthermore, relevant factors associated with HRQOL, such as socioeconomic deprivation, neurodevelopmental impairment, and prolonged hospitalizations, were not examined. Consequently, the moderate effects of these variables remain undetermined.

## Data Availability

The data that support the findings of this study are available from the corresponding author upon reasonable request.

## Acknowledgments

We would like to acknowledge Sandra Aiello and Navreet Gill, transition clinic nurses at the Labatt Family Heart Centre, for their valuable contribution in engaging patients to participate in the study.

## Sources of Funding

This research did not receive any specific grant from funding agencies in the public, commercial, or not-for-profit sectors.

## Disclosures

None

## Abbreviation

HRQOL: Health-Related Quality of Life
PedsQL^TM^: Pediatric Quality of Life Inventory
CHD: congenital heart disease
GAD: Generalized Anxiety Disorder
PHQ: Patient Health Questionnaire

## SUPPLEMENTS

**Table S1.**
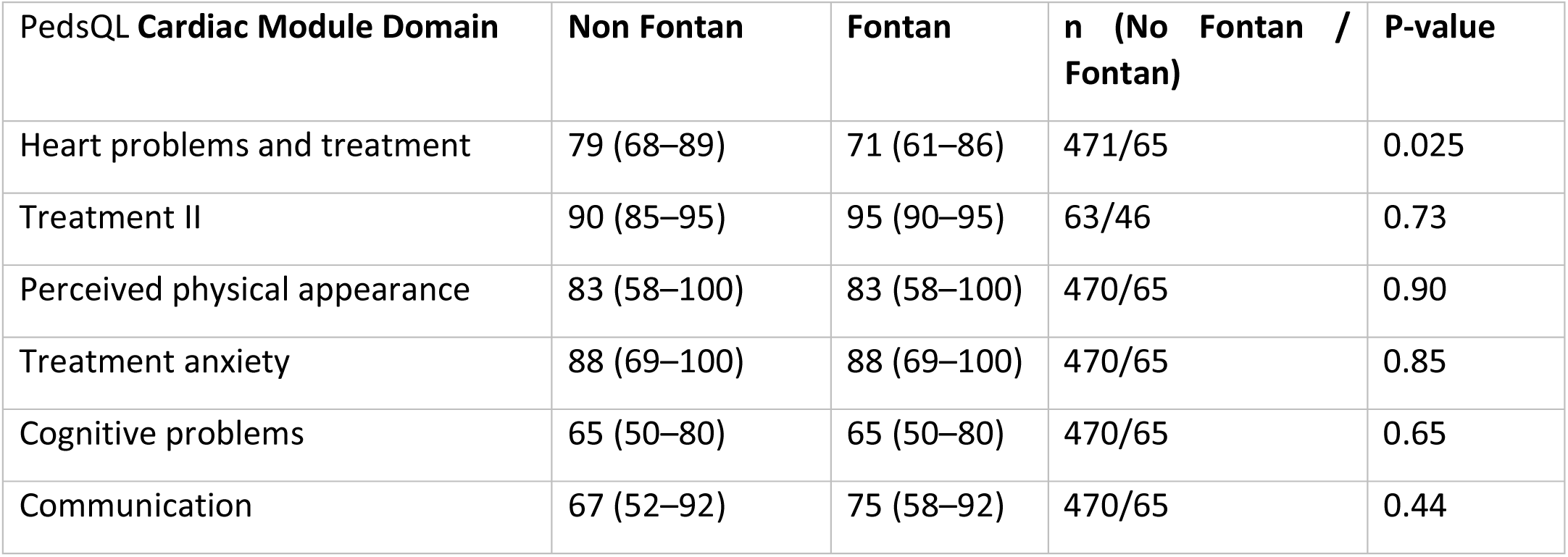
PedsQL Cardiac Module Scores by Fontan Status in Adolescents with CHD. Table S1 compares PedsQL Cardiac Module subdomains scores between adolescents with and without Fontan circulation.

| PedsQL Cardiac Module Domain | Non Fontan | Fontan | n (No Fontan / Fontan) | P-value |
| --- | --- | --- | --- | --- |
| Heart problems and treatment | 79 (68–89) | 71 (61–86) | 471/65 | 0.025 |
| Treatment II | 90 (85–95) | 95 (90–95) | 63/46 | 0.73 |
| Perceived physical appearance | 83 (58–100) | 83 (58–100) | 470/65 | 0.90 |
| Treatment anxiety | 88 (69–100) | 88 (69–100) | 470/65 | 0.85 |
| Cognitive problems | 65 (50–80) | 65 (50–80) | 470/65 | 0.65 |
| Communication | 67 (52–92) | 75 (58–92) | 470/65 | 0.44 |

**Table S2.** Transition readiness scores among adolescents with Fontan. Table S2 Transition readiness scores among adolescents with CHD, complex CHD and Fontan circulation. Data are presented as n (%) or median (IQR).

| Variable | Overall (n = 193) | Complex CHD and no Fontan (n = 128) | Fontan (n = 65) | P-value |
| --- | --- | --- | --- | --- |
| **I answer a doctor's or nurse's questions** |  |  |  | 0.60 |
| Never | 4 (2%) | 3 (2%) | 1 (2%) |  |
| Sometimes | 76 (39%) | 47 (37%) | 29 (45%) |  |
| Always | 113 (59%) | 78 (61%) | 35 (54%) |  |
| **I help to make decisions about my health** |  |  |  | 0.91 |
| Never | 11 (6%) | 8 (6%) | 3 (5%) |  |
| Sometimes | 92 (48%) | 60 (47%) | 32 (49%) |  |
| Always | 90 (47%) | 60 (47%) | 30 (46%) |  |
| **I am in charge of taking any medicine that I need** |  |  |  | 0.141 |
| Never | 25 (13%) | 19 (15%) | 6 (9%) |  |
| Sometimes | 54 (28%) | 40 (31%) | 14 (22%) |  |
| Always | 114 (59%) | 69 (54%) | 45 (69%) |  |
| **I talk to a doctor or nurse when I have health concerns** |  |  |  | 0.27 |
| Never | 19 (10%) | 13 (10%) | 6 (9%) |  |
| Sometimes | 85 (44%) | 51 (40%) | 34 (52%) |  |
| Always | 89 (46%) | 64 (50%) | 25 (38%) |  |
| **I look for an answer when I have a question about my health** |  |  |  | 0.68 |
| Never | 14 (7%) | 8 (6%) | 6 (9%) |  |
| Sometimes | 69 (36%) | 45 (35%) | 24 (37%) |  |
| Always | 110 (57%) | 75 (59%) | 35 (54%) |  |
| **I ask the doctor or nurse questions** |  |  |  | 0.093 |
| Never | 19 (10%) | 13 (10%) | 6 (9%) |  |
| Sometimes | 108 (56%) | 65 (51%) | 43 (66%) |  |
| Always | 66 (34%) | 50 (39%) | 16 (25%) |  |
| **I speak to the doctor instead of my parent(s) speaking for me** |  |  |  | 0.75 |
| Never | 30 (16%) | 19 (15%) | 11 (17%) |  |
| Sometimes | 134 (69%) | 88 (69%) | 46 (71%) |  |
| Always | 29 (15%) | 21 (16%) | 8 (12%) |  |
| **I summarize my medical history when I am asked to** |  |  |  | 0.95 |
| Never | 64 (33%) | 43 (34%) | 21 (32%) |  |
| Sometimes | 74 (38%) | 48 (38%) | 26 (40%) |  |
| Always | 55 (28%) | 37 (29%) | 18 (28%) |  |
| **I contact a doctor when I need to** |  |  |  | 0.98 |
| Never | 75 (39%) | 49 (38%) | 26 (40%) |  |
| Sometimes | 50 (26%) | 33 (26%) | 17 (26%) |  |
| Always | 68 (35%) | 46 (36%) | 22 (34%) |  |
| **I see the doctor or nurse on my own during an appointment** |  |  |  | 0.94 |
| Never | 110 (57%) | 73 (57%) | 37 (57%) |  |
| Sometimes | 69 (36%) | 45 (35%) | 24 (37%) |  |
| Always | 14 (7%) | 10 (8%) | 4 (6%) |  |
| **I drop off or pick up my prescriptions when I need medicine** |  |  |  | 0.69 |
| Never | 131 (68%) | 86 (67%) | 45 (69%) |  |
| Sometimes | 42 (22%) | 27 (21%) | 15 (23%) |  |
| Always | 20 (10%) | 15 (12%) | 5 (8%) |  |
| **I travel on my own to a doctor's appointment** |  |  |  | 0.133 |
| Never | 170 (88%) | 109 (85%) | 61 (94%) |  |
| Sometimes | 16 (8%) | 12 (9%) | 4 (6%) |  |
| Always | 7 (4%) | 7 (5%) | 0 (0%) |  |
| **I book my own doctor's appointments** |  |  |  | 0.016 |
| Never | 165 (85%) | 103 (80%) | 62 (95%) |  |
| Sometimes | 23 (12%) | 20 (16%) | 3 (5%) |  |
| Always | 5 (3%) | 5 (4%) | 0 (0%) |  |
| **Transition readiness total score** | 15 (11–17) | 15 (12–17) | 14 (11–17) | 0.57 |

**Table S3.** Correlation Between Number of Interventions and PedsQL^TM^4.0 scores. Table S3 presents the correlations between the number of prior interventions and PedsQL^TM^4.0 domains and total scores in 536 patients. Correlation coefficients and corresponding p-values are reported for each functional domain and the overall PedsQL^TM^4.0 score.

| **Variables** | **N** | **Correlation** | **P-value** |
| --- | --- | --- | --- |
| Physical function | 536 | -0.132 | 0.002 |
| Emotional function | 536 | -0.031 | 0.47 |
| Social function | 536 | -0.102 | 0.018 |
| School function | 536 | -0.047 | 0.276 |
| PedsQL™4.0 total score | 536 | -0.090 | 0.036 |

**Table S4.** PHQ-9 and GAD Scores by CHD Severity. Table S4. Summarizes the distribution of PHQ-9 depression and GAD anxiety severity categories across CHD severity groups (simple, moderate, complex) in 106 patients. Counts and percentages for each category are presented along with overall group comparisons and corresponding p-values.

| **Variable** | **Overall**<br>**(n=106)** | **Simple**<br>**(n=7)** | **Moderate**<br>**(n=62)** | **Complex**<br>**(n=37)** | **P-value** |
| --- | --- | --- | --- | --- | --- |
| **PHQ-9** |  |  |  |  | 0.84 |
| Minimal or<br>no<br>depression | 42<br>(39.6%) | 2 (28.6%) | 22 (35.5%) | 18 (48.6%) |  |
| Mild<br>depression | 33<br>(31.1%) | 3 (42.9%) | 21 (33.9%) | 9 (24.3%) |  |
| Moderate<br>depression | 18<br>(17.0%) | 2 (28.6%) | 11 (17.7%) | 5 (13.5%) |  |
| Moderately<br>severe<br>depression | 9 (8.5%) | 0 (0.0%) | 6 (9.7%) | 3 (8.1%) |  |
| Severe<br>depression | 4 (3.8%) | 0 (0.0%) | 2 (3.2%) | 2 (5.4%) |  |
| **GAD** |  |  |  |  | 0.96 |
| Minimal anxiety | 37 (34.9%) | 2 (28.6%) | 23 (37.1%) | 12 (32.4%) |  |
| Mild anxiety | 36 (34.0%) | 3 (42.9%) | 19 (30.6%) | 14 (37.8%) |  |
| Moderate anxiety | 25 (23.6%) | 2 (28.6%) | 14 (22.6%) | 9 (24.3%) |  |
| Severe anxiety | 8 (7.5%) | 0 (0.0%) | 6 (9.7%) | 2 (5.4%) |  |

**Table S5.** PHQ-9 and GAD Scores by Fontan Status. Table S5. presents the distribution of PHQ-9 depression and GAD anxiety severity categories stratified by Fontan status (Fontan vs. non-Fontan) in 106 patients. Counts and percentages for each category are reported along with corresponding p-values for group comparisons.

| Variable | No Fontan (n=93) | Fontan (n=13) | P-value |
| --- | --- | --- | --- |
| **PHQ-9** |  |  | 0.93 |
| Minimal or no depression | 35 (37.6%) | 7 (53.8%) |  |
| Mild depression | 30 (32.3%) | 3 (23.1%) |  |
| Moderate depression | 16 (17.2%) | 2 (15.4%) |  |
| Moderately severe depression | 8 (8.6%) | 1 (7.7%) |  |
| Severe depression | 4 (4.3%) | 0 (0.0%) |  |
| **GAD** |  |  | 1.00 |
| Minimal anxiety | 32 (34.4%) | 5 (38.5%) |  |
| Mild anxiety | 32 (34.4%) | 4 (30.8%) |  |
| Moderate anxiety | 22 (23.7%) | 3 (23.1%) |  |
| Severe anxiety | 7 (7.5%) | 1 (7.7%) |  |

**Table S6.** Transition clinic impact on Health-related quality of life (HRQOL) outcomes. Table S6 presents median (IQR) HRQOL domain and total scores at the first and second transition clinic visits. Corresponding p-values are provided for comparisons between visits across all domains.

| **Variables** | **First Visit**<br>**Median (IQR)** | **Second Visit**<br>**Median (IQR)** | **P value** |
| --- | --- | --- | --- |
| Physical function | 84 (75–97) | 84 (78–94) | 0.64 |
| Emotional function | 75 (56–85) | 70 (55–85) | 0.94 |
| Social function | 85 (75–100) | 85 (75–100) | 0.58 |
| School function | 65 (55–80) | 65 (50–80) | 0.68 |
| **PedsQL total score** | **77 (67–86)** | **76 (68–87)** | **0.92** |
| Heart problems and treatment | 79 (64–86) | 75 (64–86) | 0.79 |
| Treatment II | 90 (86–95) | 90 (85–95) | 0.67 |
| Perceived physical appearance | 83 (58–100) | 84 (70–100) | 0.28 |
| Treatment anxiety | 88 (69–100) | 94 (75–100) | 0.29 |
| Cognitive problems | 68 (51–81) | 65 (50–80) | 0.95 |
| Communication | 71 (50–92) | 75 (60–92) | 0.29 |

## Notes

### Competing Interest Statement

The authors have declared no competing interest.

